# Intracerebral Hemorrhage Detection in Computed Tomography Scans Through Cost-Sensitive Machine Learning

**DOI:** 10.1101/2021.10.20.21264515

**Authors:** Rushank Goyal

## Abstract

**Purpose:** Intracerebral hemorrhage is the most severe form of stroke, with a greater than 75% likelihood of death or severe disability, and half of its mortality occurs in the first 24 hours. The grave nature of intracerebral hemorrhage and the high cost of false negatives in its diagnosis are representative of many medical tasks.

**Approach:** Cost-sensitive machine learning has shown promise in various studies as a method of minimizing unwanted results. In this study, 6 machine learning models were trained on 160 computed tomography brain scans both with and without utility matrices based on penalization, an implementation of cost-sensitive learning.

**Results:** The highest-performing model was the support vector machine, which obtained an accuracy of 97.5%, sensitivity of 95% and specificity of 100% without penalization, and an accuracy of 92.5%, sensitivity of 100% and specificity of 85% with penalization, on a dataset of 40 scans. In both cases, the model outperforms a range of previous work using other techniques despite the small size of, and high heterogeneity in, the dataset.

**Conclusion:** Utility matrices demonstrate strong potential for sensitive yet accurate artificial intelligence techniques in medical contexts and workflows where a reduction of false negatives is crucial.

## 1 Introduction

Intracerebral hemorrhage (ICH) is a neurological condition occurring due to the rupture of blood vessels in the brain parenchyma [1]. is the most severe form of stroke, with the chance of death or severe disability exceeding 75% and only 20% of survivors remaining capable of living independently after 1 month [2, 3]. It has an incidence of 24.6 per 100,000 person-years and accounts for 10-15% of all strokes [4]. An early diagnosis of ICH is also crucial as half of the mortality occurs in the first 24 hours [5]. Computed tomography (CT) scans are currently the preferred non-invasive approach for ICH detection [6]. The diagnosis time for ICH remains very long, reaching 512 minutes in one study, which, coupled with a high misdiagnosis rate of 13.6% as one estimate calculated, makes it a prime candidate for workflow improvement through machine learning [5, 6].

The conditions and characteristics that are specific to medicine must be taken into consideration during the application of machine learning techniques to medical problems such as ICH diagnosis. A primary concern is that false negatives are usually much more costly than false positives [7]. The frequent under-representation of the minority class coupled with the increased emphasis on its correct prediction makes this a challenging problem for artificial intelligence [8].

Taking the disproportionate costs of false negatives into account (more broadly, a technique called cost-sensitive learning) has resulted in positive outcomes in other medical tasks [9–15]

However, there is very little literature on the application of such techniques in hemorrhage diagnosis. Hence, the aim of this research was to test the results of the implementation of cost-sensitive learning (specifically a utility matrix) on ICH classification.

## 2 Materials and Methods

The data used for the study were obtained under a CC0: Public Domain license, released by Kitamura [16]. The dataset consisted of 200 anonymized, publicly-available images of non-contrast computed tomography (CT) scans (brain window), 100 of which contained instances of intraparenchymal hemorrhage with or without intraventricular extension, and 100 of which did not. A sample of 4 images from the data is shown in Figure 1. Figure 1a and 1b show scans without intracerebral hemorrhage, at the level of the lateral ventricles and third ventricle, respectively. Figure 1c displays a large intracerebral hemorrhage with intraventricular extension. Figure 1d contains a small intracerebral hemorrhage without intraventricular extension.

**Figure 1:**
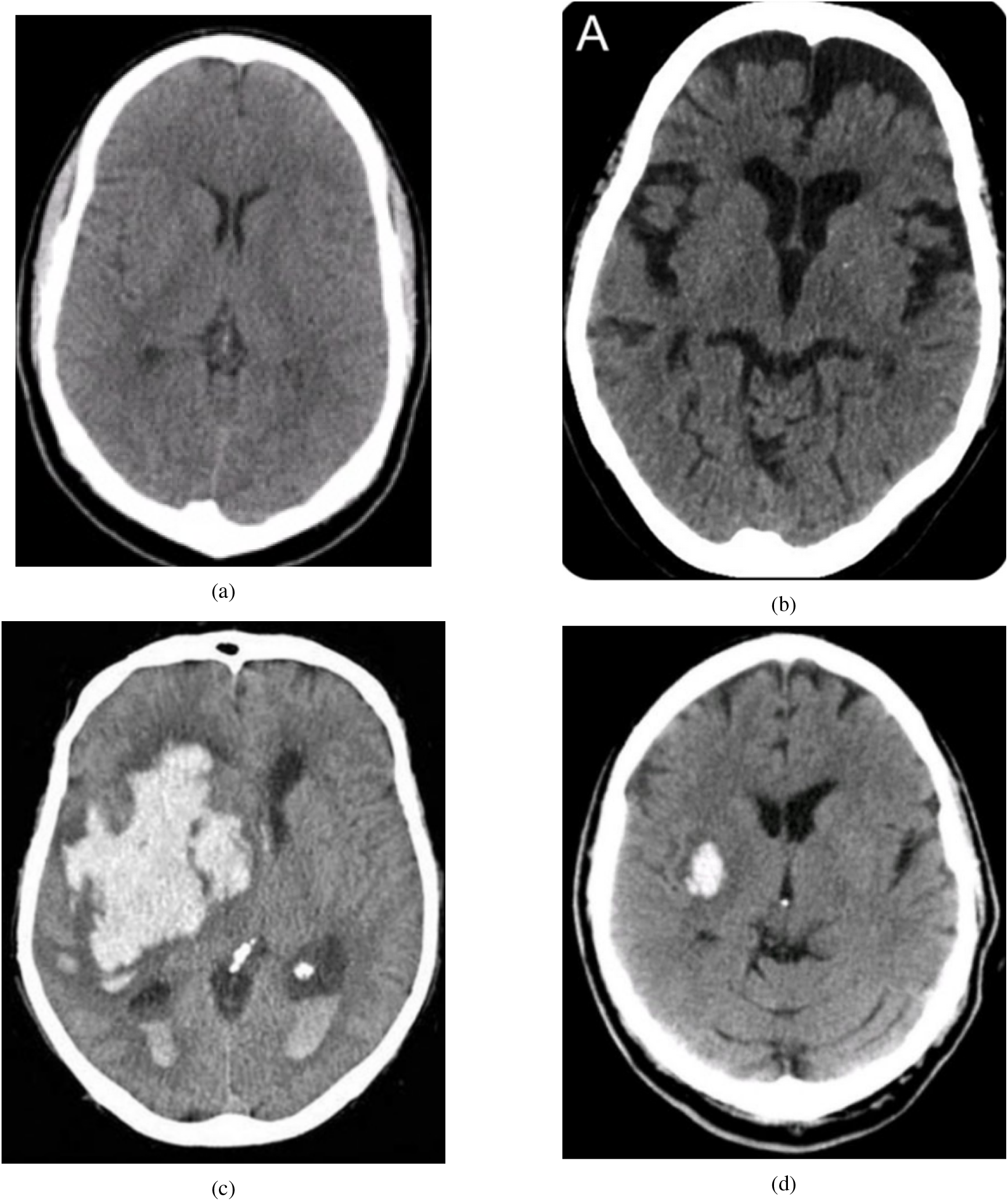
Sample Dataset Images

A trained radiologist confirmed the veracity of these images, and was unable to find any mislabeled images. Thus none of the images was discarded. To obtain the most accurate representation of model performance in real-life clinical scenarios, the images were not augmented in any way. Two datasets were subsequently created: a training dataset with 160 images and a testing dataset with 40 images. Both had equal numbers of hemorrhagic and non-hemorrhagic CT scans. It should be noted that the dataset consisted of images taken from searches on the World Wide Web, hence introducing a high level of heterogeneity due to variations in source machines, patient conditions, scan time, radiation dose, etc. This problem is compounded by the small dataset size, hence the results obtained here are likely to only be conservative estimates of the real potential of the techniques employed [17, 18].

6 machine learning models were chosen for the study. The specific parameter configurations for these models are provided in Table 1, and are important for reproducibility. A random seed of 0 was used in each case. The models were trained using Wolfram Mathematica Desktop Version 12.3.0, making use of the Classify[] function [19].

**Table 1:** Parameter Specifications

| Name | Parameters |
| --- | --- |
| Random Forest | FeatureFraction = $\frac{1}{4\sqrt{10}}$, LeafSize = 5, TreeNumber = 50, DistributionSmoothing = 0.5 |
| Decision Tree | DistributionSmoothing = 1, FeatureFraction = 1 |
| Gradient Boosted Trees | BoostingMethod = Gradient, MaxTrainingRounds = 50, LeavesNumber = 13, LearningRate = 0.1, MaxDepth = 6, LeafSize = 15, L1Regularization = 0, L2Regularization = 0 |
| Nearest Neighbors | NeighborsNumber = 5, DistributionSmoothing = 0.5, NearestMethod = Scan |
| Support Vector Machine | KernelType = RadialBasisFunction, GammaScalingParameter = 0.00113124, PolynomialDegree = 3, SoftMarginParameter = 1, BiasParameter = 1, MulticlassStrategy = OneVersusOne |
| Logistic Regression | L1Regularization = 0, L2Regularization = 100, OptimizationMethod = LBFGS |

### 2.1 Decision Tree

A decision tree is a machine learning model that consists of nodes and branches, and is built using a combination of splitting, stopping and pruning [20]. Combining all of these techniques, while not guaranteed to result in the theoretically-optimal decision tree – as that would require an exceedingly long time – still results in a highly useful model [21].

#### 2.1.1 Nodes

The root node is the first type of note, and it represents a decision that divides the entire data into two or more subsets. Internal nodes represent more choices that can be used to further split subsets. Eventually, they end in leaf nodes, representing the final result of a series of choices. Any node emanating from another node can be referred to as its child node [20].

#### 2.1.2 Branches

Branches are representations of the outcomes of decisions made by nodes and connect them to their child nodes [20].

#### 2.1.3 Splitting

Both discrete or continuous variables can be used by decision trees to set criteria for different nodes, either internal or root, that are used to split the data into multiple internal or leaf nodes. Decision trees choose between different splitting possibilities using certain measures of the child nodes, such as entropy, Gini index, and information gain, to optimize the splitting choices [20].

#### 2.1.4 Stopping

If allowed to split indefinitely, a decision tree model could achieve 100 % accuracy simply by splitting over and over until each leaf node only had one data sample. However, such a model would be vastly overfitted and would not generalize well to test data. Thus there needs to be a limit (often in the form of maximum depth allowed or minimum size of leaf nodes) [20].

#### 2.1.5 Pruning

There are two types of pruning. Pre-pruning uses multiple-comparison tests to stop the creation of branches that are not statistically significant, whereas post-pruning removes branches from a fully-generated decision tree in a way that increases accuracy on the validation set, which is a special subset of the training data not shown to the model while training [20].

### 2.2 Random Forest

A random forest, as the name suggests, is a collection of randomized decision trees that is suitable for situations where a simple decision tree could not capture the complexity of the task at hand [22]. The random forest model performs a ‘bootstrap’ by choosing *n* times from *n* decision trees with replacement [23]. The averaging of these predictions is called bagging (short for bootstrap-aggregating) and is a simple way to improve the performance of weak models, which many decision trees are when the task is complex [24].

### 2.3 Gradient Boosted Trees

Gradient tree boosting constructs an additive model, using decision trees as weak learners to aggregate, similar to the concept of random forests, but it does so sequentially; stochastic gradient descent is then used to minimize a given loss function and optimize the construction of the model as more trees are added [25, 26].

### 2.4 Nearest Neighbors

The nearest neighbors model is based on the concept that the closest patterns of the data sample in question offer useful information about its classification. Thus the model works by assigning each data point the label of its *k* closest neighbors, where *k* is specified manually [27].

### 2.5 Support Vector Machine

Support vector machines are machine learning models for classification problems [28].

#### 2.5.1 Separating Hyperplane

For *n*-dimensional data, a hyperplane (straight line in a higher-dimensional space) of *n-1* dimensions is used to separate the data into two different groups such that the distance from the clusters is maximized and the hyperplane is ‘in the middle’, so to speak. This hyperplane also dictates which labels will be assigned to the samples from the test set [28].

#### 2.5.2 Soft Margin

Most datasets, of course, do not have clean boundaries separating different clusters of samples. There will also be outliers, and an optimal model would allow for a certain number of outliers to avoid overfitting while also limiting misclassifications. The soft margin roughly controls the number of examples allowed on the wrong side of the hyperplane as well as their distance from the hyperplane [28].

#### 2.5.3 Kernel Function

A kernel function refers to one or more mathematical operations that project low-dimensional data to a higher-dimensional space called a feature space, with the goal being the easier separation of the example data [29].

### 2.6 Logistic Regression

Logistic regression is a mathematical model that describes the relationship of a number of features (*X*_1_, *X*_2_, …, *X*_*k*_) to a dichotomous result. It makes use of the logistic function, given below [30].

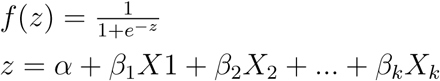

For the second part of the study, the concept of utility functions was used. A utility function is a mathematical function through which preferences for various outcomes can be quantified [31]. In the case of a utility matrix for classification problems, *U*_*ij*_ provides the utility where *i* is the ground truth and *j* is the model prediction [32]. It is also referred to as a cost matrix [33].

The utility matrix created for this study is shown in Table 2. The sole difference from the default utility matrix is that false negatives (Row 1, Column 2) now have a utility of -*x* instead of 0. Subsequent experimentation with different values of *x* was done and the results were recorded.

**Table 2:** Utility Matrix

|  |  |  |
| --- | --- | --- |
|  | Hemorrhage | No Hemorrhage |
| Hemorrhage | 1 | $-x$ |
| No Hemorrhage | 0 | 1 |

Essentially, the models trained using this matrix will have an aversion to false negatives, thus decreasing the number of false negatives at the expense of potentially increasing the number of false positives. The strength of this aversion and subsequent change is quantified by *x*. A certain amount of trial and error is needed since, for most medical problems, the cost is not certain and needs to be estimated [34]. The cost is context-specific; depending on the workflow where the algorithm is being implemented, the goals for sensitivity and specificity will vary and the cost must be determined accordingly.

In Round 2, the models were re-trained with the same parameters as the previous set, with the only difference being that the value for the ‘UtilityFunction’ parameter was set to the utility matrix provided in Table 2.

## 3 Results

The final results for all of the models trained are given in Table 3. The probability histograms for the 3 most accurate models are given in Figure 2. These histograms show the actual-class probability of the 40 test samples. Table 4 lists the results for each of the models after setting their utility function to the matrix given in Table 2, for three values of −*x*. Finally, Figure 3 shows, for the top three models, how their sensitivities and specificities change as the penalty is increased, with 0 being the default penalty that corresponds to Table 3.

**Table 3:** Round 1 Results

|  | Accuracy | Sensitivity | Specificity |
| --- | --- | --- | --- |
| Decision Tree | 0.600 | 0.800 | 0.400 |
| Random Forest | 0.700 | 0.750 | 0.650 |
| Gradient Boosted Trees | 0.900 | 0.900 | 0.900 |
| Nearest Neighbors | 0.825 | 0.700 | 0.950 |
| Support Vector Machine | 0.975 | 0.950 | 1.00 |
| Logistic Regression | 0.925 | 0.900 | 0.950 |

**Table 4:** Round 2 Results

|  | Penalty of -1 |  |  | Penalty of -2 |  |  | Penalty of -3 |  |  |
| --- | --- | --- | --- | --- | --- | --- | --- | --- | --- |
|  | Acc | Sens | Spec | Acc | Sens | Spec | Acc | Sens | Spec |
| Decision Tree | 0.500 | 1.00 | 0 | 0.500 | 1.00 | 0 | 0.500 | 1.00 | 0 |
| Random Forest | 0.650 | 0.900 | 0.400 | 0.650 | 0.900 | 0.400 | 0.700 | 1.00 | 0.400 |
| Gradient Boosted Trees | 0.900 | 1.00 | 0.800 | 0.900 | 1.00 | 0.800 | 0.850 | 1.00 | 0.700 |
| Nearest Neighbors | 0.800 | 0.900 | 0.700 | 0.800 | 0.900 | 0.700 | 0.625 | 0.950 | 0.300 |
| Support Vector Machine | 0.925 | 1.00 | 0.850 | 0.925 | 1.00 | 0.850 | 0.900 | 1.00 | 0.800 |
| Logistic Regression | 0.925 | 0.950 | 0.900 | 0.900 | 0.950 | 0.850 | 0.850 | 1.00 | 0.700 |

**Figure 2:**
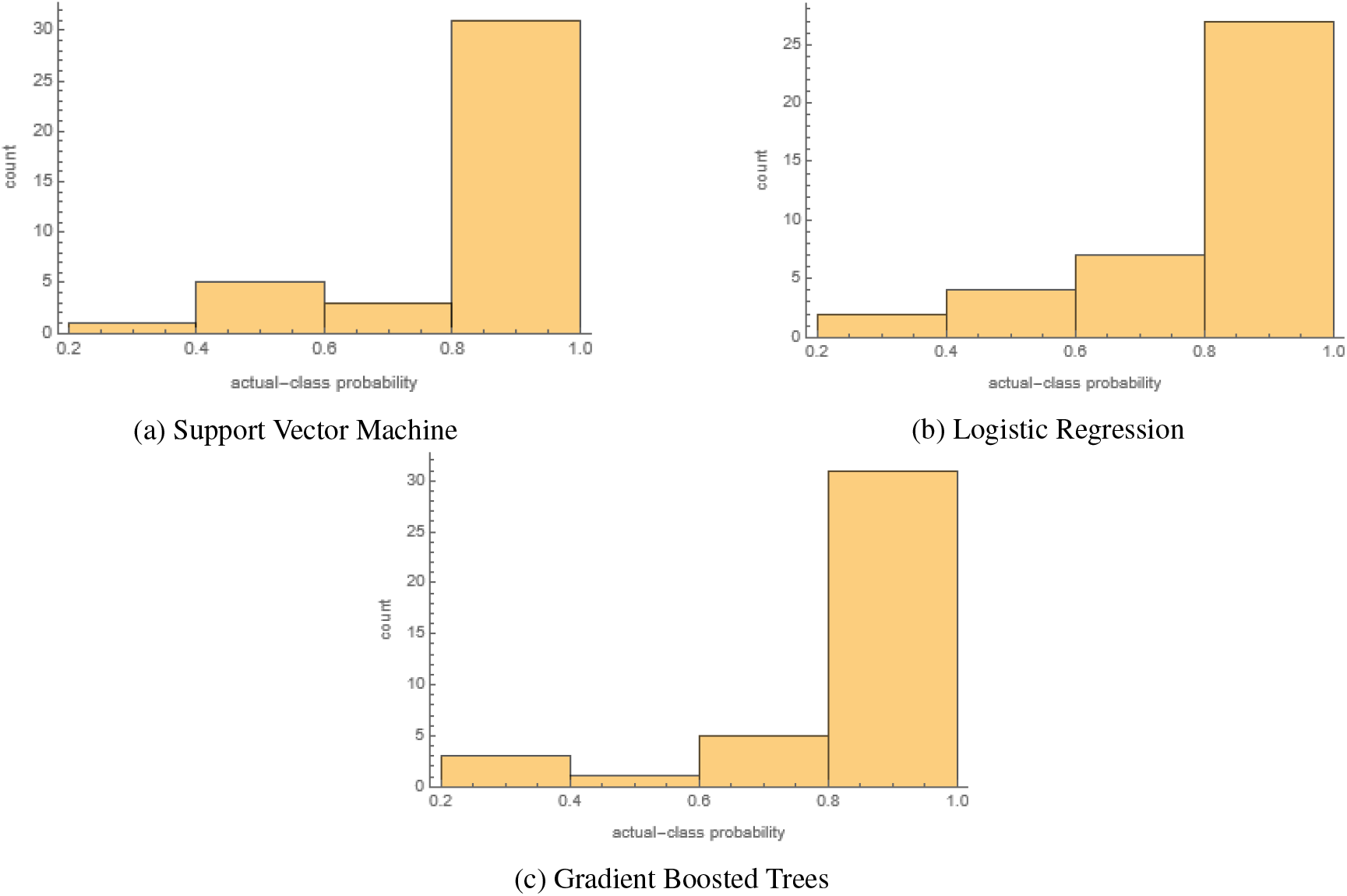
Probability Histograms

**Figure 3:**
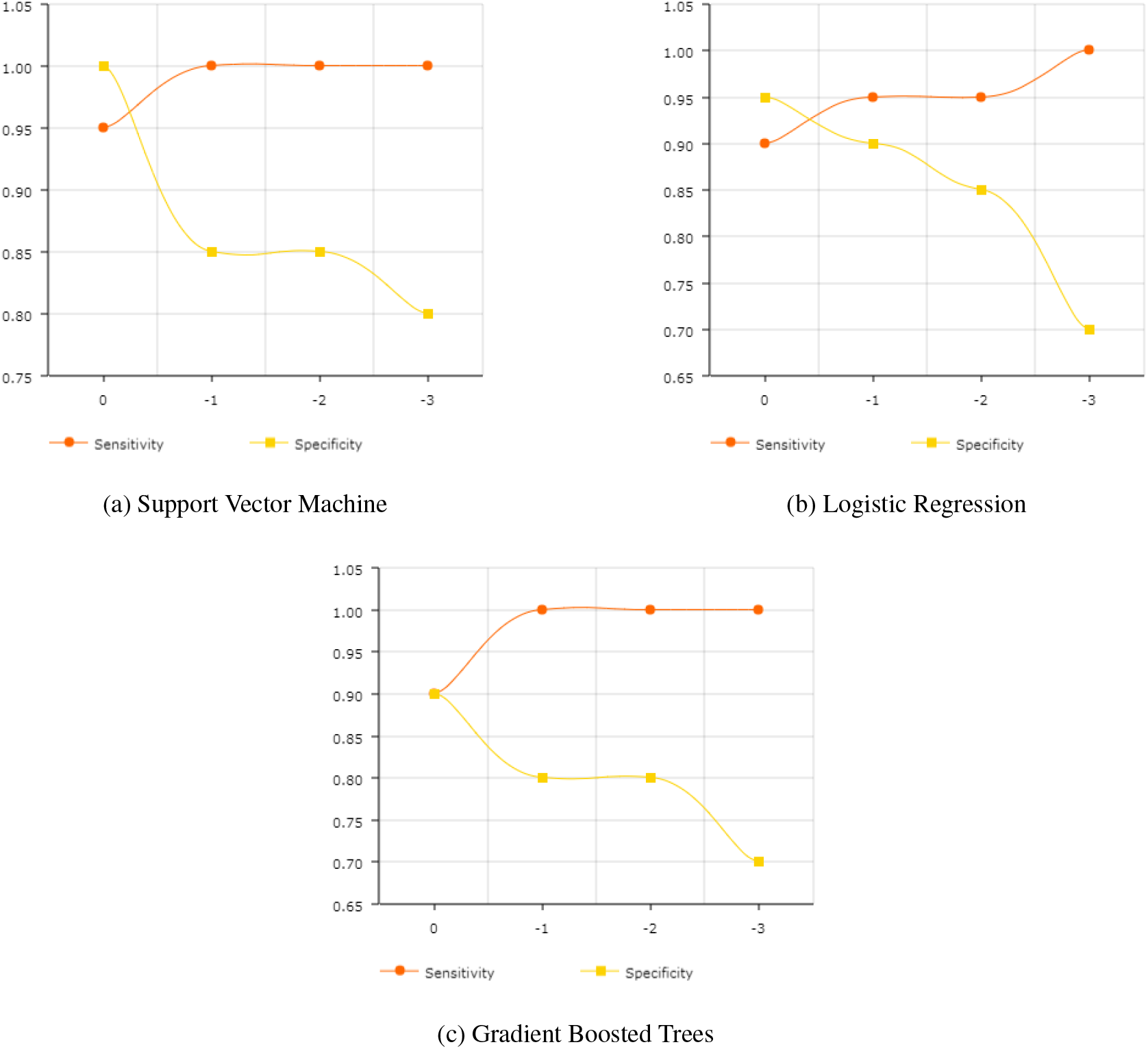
Trends for Sensitivites and Specificities By Penalty

According to the data in Table 3, the support vector machine model is the most accurate as well as the most sensitive and specific. Logistic regression and gradient boosted trees score second and third respectively in overall accuracy, though the nearest neighbors model has a higher specificity than gradient boosted trees, coming in second and tied with logistic regression.

Interestingly, while Table 3 shows that the support vector machine is more accurate overall, logistic regression actually appears to be more certain in its predictions, as demonstrated by the difference in the number of samples in the 0.4-0.6 bin. Overall, though, all three of the best-performing models seem highly certain in the correct predictions that they are making, as the vast majority of samples fall in the 0.8-1.0 bin.

In Table 4, for a penalty of -1, three models give a 100% sensitivity, although for the decision tree that comes at the cost of having a 0% specificity. The support vector machine performs the best, matching the 100% sensitivity of gradient boosted trees. Its specificity of 85% is less than that achieved by logistic regression, though its sensitivity is greater.

The results for a -2 penalty are almost exactly the same, with the only difference being a slightly reduced accuracy for logistic regression.

When the penalty is increased to -3, model performances decrease along the board, except for random forest, which performs better). While the sensitivities for almost all the models are now 100%, they come at the expense of drastically reduced specificities.

Further values of −*x* were not tested as most model performances had started rapidly deteriorating at a -3 penalty. The random forest, however, might perform better at more negative penalties, based on the observed trends.

As expected, Figure 3 shows that greater penalties yield reductions in specificity and growth in sensitivity. Though in cases like Figure 3a and Figure 3c, where the sensitivity maxes out early, additional penalties only decrease the specificity. While tweaking the penalties, some changes, such as the one 0.85 to 0.70 in the specificity in Figure 3b, can be quite drastic, making it all the more important to conduct detailed experimentation while determining the penalty.

## 4 Discussion

The Round 1 results compare favorably to prior studies conducted for ICH identification, especially taking into the account the small training set and heterogeneity in the training data. For instance, work done by Majumdar et al. [35] shows a sensitivity of 81% and specificity of 98%. In one study, the model accuracy of 82%, recall of 89% and precision of 81% remarkably still resulted in a better recall than 2 of the 3 senior radiologists used for benchmarking [36]. More recent research by Dawud et al. [37] resulted in the development of three models, with accuracies of 90%, 92% and 93%. Jnawali et al. [38] performed a study with dataset heterogeneity comparable to ours, although with a considerably larger dataset size of 1.5 million images, and achieved an AUC score of 0.87. A study using 37,000 training samples, also with pronounced heterogeneity in the dataset, obtained an accuracy of 84%, a sensitivity of 70%, and a specificity of 87% [5]. There were also two studies that achieved better accuracies of 99% and one study with an AUC of 0.991 [39–41].

As for cost-sensitive machine learning, only one example could be found for the application of such a technique on intracerebral hemorrhage in prior literature, where the authors ended up achieving an overall sensitivity of 96% and specificity of 95% on a training dataset of 904 cases; the cost in this case was introduced through a modified loss function instead of a utility matrix (used in this study) [42]. A smaller training dataset of 160 images combined with greater heterogeneity likely reduced model performances in this study compared to Lee et al.’s work [17, 18, 42].

The accuracy of the cost-sensitive support vector machine exceeds a number of prior studies’ performances. Comparing the accuracy against the array of research mentioned previously, it manages an accuracy greater than the models constructed by Dawud et al., Grewal et al., Arbabshirani et al. and Jnawali et al., while still maintaining a 100% sensitivity [5, 36–38]. As Rane and Warhade pointed out in their literature review, a cost-sensitive algorithm shows great potential as a strategy for more effective diagnosis, and the results confirm this, with the cost-sensitive models demonstrated here outperforming a number of other techniques [43].

### Applications

Considering the details of utility matrix application and the results it produces, such a type of optimization is suited for a workflow where the artificial intelligence is not working in isolation. Since penalization for false negatives can end up reducing specificities, a further test might become necessary, whether it is a separate algorithm, human clinician(s), or something else. Cost-sensitive learning is ideal in situations where misdiagnosing a positive case could have serious negative impacts, but at the same time the misdiagnosis of a negative case would not cause much inconvenience.

### Limitations and Future Research

Two main limitations of this study were, as mentioned, the small sample size and high heterogeneity within the dataset. Moreover, due to the nature of the dataset, distinctions based on sex, ethnicity or age weren’t possible. Future research that applies cost-sensitive techniques to larger, more standardized datasets, as well as utilizing datasets with details about sex, age, ethicity, etc., could offer greater insights into the advantages and drawbacks of utility matrix-based artificial intelligence algorithms.

### Conclusion

Clearly, utility matrices have potential as a tool for minimizing unwanted false negatives while still providing accurate overall results. There is a trade-off between sensitivity and specificity, and a suitable value must be chosen after multiple trial and error estimates, but once the ideal penalty has been determined, a cost-sensitive model achieves the given goal better than a more general technique. More work needs to be done on a variety of use cases, not just intracerebral hemorrhage detection, to test the performances of such techniques and their viability in clinical scenarios.

## Data Availability

The data used in the manuscript are available under a CC0: Public Domain license at https://www.kaggle.com/felipekitamura/head-ct-hemorrhage/.

## 5 Data Availability Statement

The dataset employed was provided under a CC0: Public Domain license by Kitamura [16]. The code for the study can be accessed at https://github.com/rushankgoyal/cost-sensitive-hemorrhage

## 7 Ethical Declarations

This research study was conducted retrospectively using human subject data made available in open access. Ethical approval was not required as confirmed by the institutional review board (IRB) of the Sri Aurobindo Institute of Medical Sciences in Indore, India.

